# Single-cell atlas of intracranial arteries reveals sex-specific gene regulation linked to intracranial aneurysm risk

**DOI:** 10.1101/2025.11.06.25339724

**Authors:** Dennis P. Maas, Ynte M. Ruigrok, Mark K. Bakker

## Abstract

**BACKGROUND:** Rupture of an intracranial aneurysm (IA) results in aneurysmal subarachnoid hemorrhage, a severe form of stroke that is more prevalent in women than in men. The biological mechanisms underlying IA formation and the observed sex differences in prevalence in women remain poorly understood. Here, we present a single-cell RNA sequencing (scRNA-seq) atlas of intracranial arteries and use it to analyze sex-specific differences in gene expression and cell type composition. We further apply this atlas as a reference for deconvolution of IA bulk RNA-seq samples to study the effect of sex- and rupture status on cell type composition.

**METHODS:** scRNA-seq was performed on human intracranial artery samples (n=7) from the circle of Willis without IAs, yielding high-quality transcriptomic profiles from 55,371 cells. We characterized cell types and examined sex differences in gene expression and cell type proportions. Using this dataset as a reference, we performed bulk RNA-seq deconvolution on IA samples to assess differences in cell type proportions associated with sex or rupture status.

**RESULTS:** We identified seven distinct cell types, as well as multiple subpopulations within structural arterial cell types. While cell type proportions were comparable between male (n=3) and female (n=4) arterial samples and IA samples after RNA deconvolution, multiple biological pathways showed sex-specific regulation. Also, ruptured IAs had higher proportions of myeloid lineage cells compared to unruptured IAs.

**CONCLUSIONS:** Analysis of our publicly available single-cell atlas of intracranial arteries revealed no substantial sex differences in cell-type composition. However, pronounced sex-specific gene expression differences within the structural arterial cell types were observed, which may contribute to the higher prevalence of IAs in women. This single-cell atlas provides a valuable resource for further research into the pathogenesis of IAs, as well as other diseases of the intracranial arteries.

## Introduction

The circle of Willis (CoW) is a network of intracranial arteries supplying blood to the brain.^1^ Arterial bifurcations within the CoW are common sites of intracranial aneurysm (IA) formation, affecting ∼3% of the general population, with two-thirds of whom are women.^2^ In some cases, the IA ruptures, causing an aneurysmal subarachnoid hemorrhage, a severe type of stroke that is fatal in one-third of patients and disabling in another third.^3^ Shear stress and inflammation are implicated in IA formation^4^, but the molecular mechanisms and the contributions of specific cell types of the arterial wall remain poorly understood. It is also unclear whether these molecular mechanisms differ between sexes, which could underlie the higher risk of IAs in women.

Single-cell RNA sequencing (scRNA-seq) is a powerful technique for uncovering the molecular mechanisms underlying disease and enables cell type-specific analyses of gene expression, including sex-specific effects. While prior studies have used scRNA-seq to a limited number of IA samples^5–7^, they lacked healthy intracranial arteries as controls. The intracranial arteries of the CoW differ from extracranial arteries by lacking a lamina externa^8^, suggesting that their cells may also exhibit a distinct gene expression pattern. Therefore, scRNA-seq data from the CoW can provide valuable insights into the molecular mechanisms and cell types critical to IA formation and rupture.

In this study, we present the first scRNA-seq atlas of the intracranial arteries from the CoW and use it to analyze sex-specific differences in cell type composition and gene expression patterns within structural arterial cell types. We further apply this dataset for bulk RNA-seq deconvolution of IA samples^9^ to uncover differences in cell type proportions between men and women, and between ruptured and unruptured IAs.

## Methods

### Data availability

Our scRNA-seq atlas will be made publicly available upon publication. In addition, raw sequencing data, count matrixes and cell metadata will be made available in the Gene Expression Omnibus database.

### Sample acquisition

Human CoW samples (n=7) were obtained from brains of donors without dementia and without IAs through the Netherlands Brain Bank. The entire CoW was carefully dissected between 4 to 6.5 hours postmortem, snap-frozen, and stored in saline at -80°C to preserve tissue integrity for analyses. Donor characteristics are described in Table S1.

### Sample processing

Cell dissociation, scRNA-seq library preparation, and sequencing were performed by Single Cell Discoveries (Utrecht, Netherlands). Tissues were processed using the “Tissue Fixation & Dissociation for Chromium Fixed RNA Profiling” protocol from 10x Genomics (CG000553). scRNA-seq libraries were prepared using the “Chromium Fixed RNA Profiling Reagent Kits for Multiplexed Samples” protocol from 10x Genomics (CG000527). Sequencing was performed on an Illumina NovaSeq X Plus using 2 × 50 bp, 10B flowcells, following the manufacturer’s instructions and the 10x Genomics guidelines for library loading and sequencing configuration.

### scRNA-seq data processing

Raw FASTQ files from scRNA-seq were mapped to the GRCh38-2020-A human genome, and gene counts were generated using 10x Genomics Cell Ranger v7.2.0. The Cell Ranger multi pipeline with the v1.0.1 probe set was used to process the samples after demultiplexing. Cell calling and ambient RNA removal were performed per sample using CellBender v0.3.0^10^, with 250 epochs and a learning rate of 1.25 × 10⁻⁵, both optimized based on preliminary runs. All clustering analyses were performed using Seurat v5.1.0^11^ in R v4.3.3. Initial filtering steps included exclusion of cells expressing fewer than 200 genes, and removing genes detected in fewer than 5 cells. Doublet identification was performed using the scDblFinder v1.16.0^12^ package, which requires preliminary clustering for each sample individually. For this preliminary clustering, we applied default log-normalization, variable feature selection, scaling and principal component analysis. Clustering was performed using the Leiden algorithm v0.10.2^13^ with the igraph method v0.10.12^14^ and using the first 15 principal components. Resolutions ranging from 0.05 to 1.0 in 0.05 increments were tested were used to find the optimal clustering resolution, which was selected based on the highest mean silhouette score from the approxSilhouette() function in the bluster v1.12.0^15^ package.

For the final clustering, only singlets were loaded into a new Seurat object with each sample in a separate layer. Cell quality control metrics were added with the Add_Cell_QC_Metrics() function from the scCustomize v2.1.2^16^ package. The second filtering step removed cells with fewer than 400 genes, more that 5% mitochondrial genes or more than 2% hemoglobin genes. Last, genes expressed in fewer than 5 cells were also removed. Default log-normalization, variable feature selection, and principal component analysis were used. To mitigate the effect of sequencing depth on clustering, the number of unique molecular identifiers (UMIs) was regressed out during scaling. To integrate the different samples, the RPCAIntegration method in the IntegrateLayers() function was used with default parameters. The Leiden algorithm was used in the same way as in the preliminary clustering. The manifold approximation and projection (UMAP) algorithm in the RunUMAP function, also using the first 15 principal components, was used to visualize the clusters. Clustering resolutions were determined based on mean silhouette scores, gene set enrichment analysis results, and UMAP visualisations. We used a resolution of 0.1 for our level 1 clustering, which identified the main categories of structural arterial cell types, including vascular smooth muscle cells (vSMCs), perivascular fibroblasts (PVFs), and vascular endothelial cells (vECs). Next, our level 2 clustering involved subclustering of these structural arterial cell types. For each cell type, all cells from their corresponding level 1 cluster were selected, and the same clustering pipeline was applied. For vECs, the integration parameter k.weight was set to 60, as the cluster contained too few cells per sample for the integration to run with default parameters. The clustering resolutions were 0.25 for vSMCs, 0.25 for PVFs, and 0.2 for vECs.

### Differential cell type proportion analysis

Differences in cell type proportions in the scRNA-seq data between male and female samples were analyzed using the propeller method from the speckle package v1.4.0^17^ and Seurat object containing raw cell type counts as input. Proportions were transformed using the arcsine (‘asin’) transformation. Cell type proportion differences were assessed for each cell type separately using a two-tailed t-test and adjusted for multiple testing with the Benjamini-Hochberg false discovery rate (FDR) method.

### Differential expression analysis

Differential expression analysis was performed using generalized linear models implemented in DESeq2 v1.42.0^18^ to compare gene expression between clusters or between sexes. Patient age was centered and scaled using the base R scale() function and included as a covariate. To ensure compatibility with DESeq2, pseudobulk objects were generated using Seurat’s AggregateExpression() function, grouped by sample ID, scaled age and the comparison variable. When analyzing sex effects, Y chromosome genes were excluded when studying the effect of sex.

### Pathway analysis

Gene set enrichment analysis was performed in R using the clusterProfiler v4.10.0^19^ package. Genes were ranked according to the Wald test statistic from DESeq2. The org.Hs.eg.db v3.18.0 package was used as the species gene database. Gene Ontology (GO) enrichment was tested with the gseGO() function. The hallmark gene set collection was obtained via the msigdbr package v7.5.1^20^ and tested for enrichment with the GSEA() function. P-values were adjusted for multiple testing with the Benjamini-Hochberg FDR method.

### Deconvolution analysis

Cell type deconvolution was used to estimate cell type proportions of bulk RNA-seq data, from 44 IA samples generated in a previous project (accession number GSE122897^9^), using our level 1 scRNA-seq atlas (i.e. main categories of structural arterial cell types) as reference. Deconvolution analysis was carried out using BayesPrism v2.1.1^21^ with default parameters. Genes located on chromosomes X and Y, as well as mitochondrial and ribosomal genes, and those expressed in fewer than five cells were excluded. After this, only protein coding genes were selected for the analysis. BayesPrism updates cell type proportions by pooling information across samples, which could introduce a confounding effect. To avoid this, we used the “initial” cell type proportions from BayesPrism, which are estimated independently for each sample. Structural arterial cell type proportions were calculated by excluding non-structural cell types and rescaling the remaining proportions to sum to 100%. To study sex effects, 13 male and 31 female samples were compared. To assess the effect of rupture status, 21 unruptured IAs were compared to 22 ruptured IAs; one sample with unknown rupture status was excluded from this analysis. Statistical analysis was performed using the same framework as for the differential cell type proportion analysis of the scRNA-seq data. For the propeller method, data was processed using the convertDataToList() function with default parameters.

## Results

### Single-cell RNA sequencing atlas of the CoW

After quality control of sequencing data from seven human CoW samples (Figure 1A), high quality gene expression profiles were obtained from 55,371 cells (Figure S1). The level 1 clustering identified seven distinct cell type clusters: the main structural arterial cell types, which are vSMCs, PVFs, and vECs, together with meningeal fibroblasts (MFs), glial cells, myeloid lineage cells and lymphoid lineage cells (Figure 1; Table S2 and S3). vSMCs, expressing ACTA2 and MYH11, were the most abundant cell type (Figure S1I). PVFs were enriched for extracellular matrix-related genes, including LAMA2 and COL4A4, whereas MFs were enriched for membrane transport-related genes, such as SLC4A4 and KCNMA1. vECs expressed typical endothelial markers, such as VWF and PECAM1. The glial cluster expressed genes related to myelination, including MPZ and PLP1, as well as the astrocyte marker GFAP, suggesting a heterogeneous population that may include Schwann cells, oligodendrocytes, and astrocytes. Two immune cell populations were identified: a myeloid lineage cluster, expressing CD163 and CD14, and a lymphoid lineage cluster, expressing IL7R and KLRK1.

**Figure 1.**
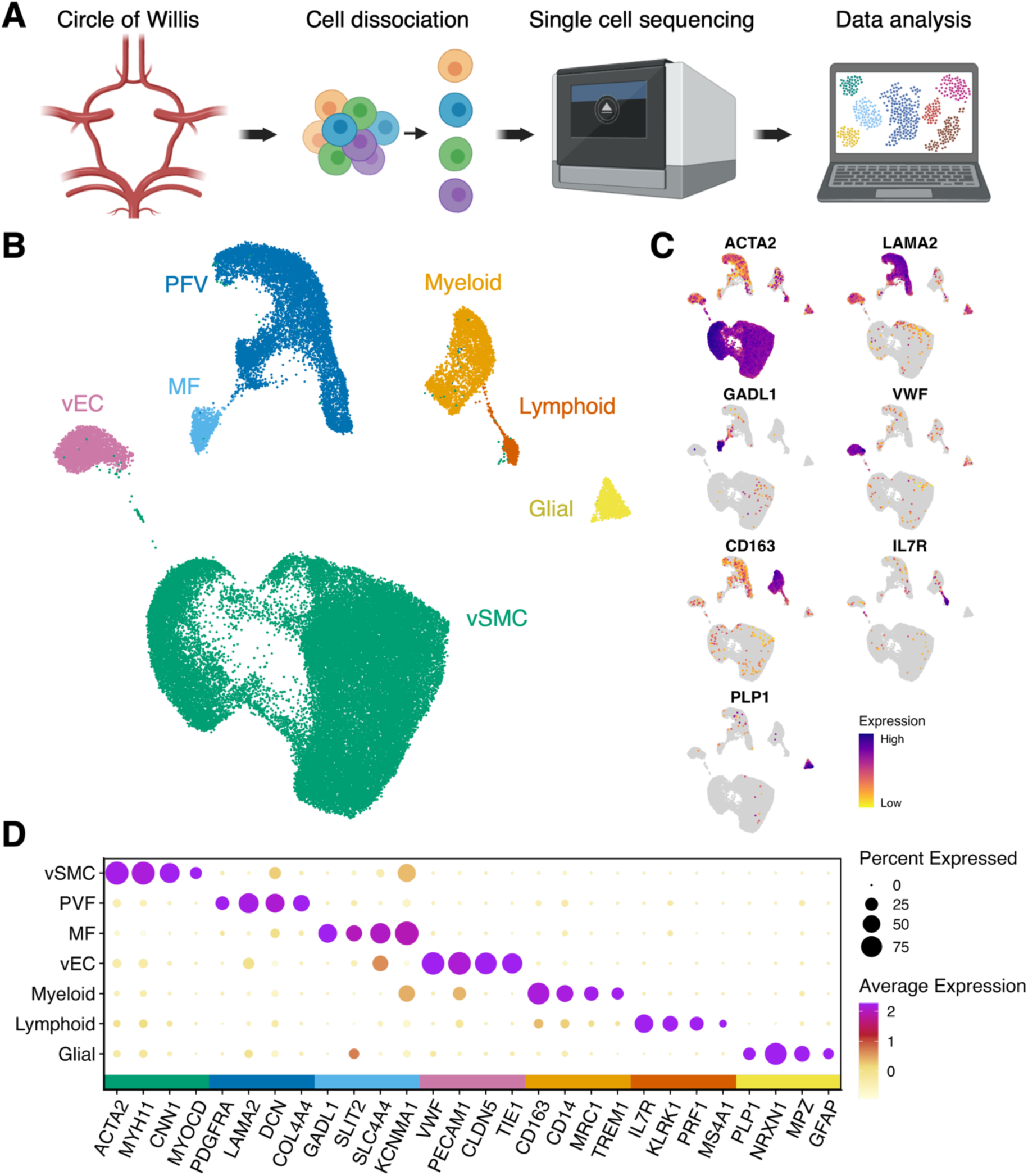
Single-cell RNA sequencing (scRNA-seq) atlas of the intracranial arteries of the circle of Willis (CoW). (**A**) Schematic overview of the research design^34^. (**B**) Uniform Manifold Approximation and Projection (UMAP) visualization of the seven cell types in the CoW arteries. (**C**) UMAP visualization showing gene expression of key cell type markers. (**D**) Dot Plot showing the expression of marker genes for the different cell types. MF, meningeal fibroblast; PVF, perivascular fibroblast; vEC, vascular endothelial cell; vSMC, vascular smooth muscle cell.

### Level 2 clustering uncovers granular structural arterial cell types

To further characterize the cell types of the CoW arteries, we performed additional clustering of the structural cell types of the arterial wall, vSMCs, PVFs and vECs (Figure 2; Table S4 and S5). This level 2 clustering revealed multiple subpopulations within each level 1 cell type. In the vSMCs, five subclusters were identified, which were designated vSMC1-5 (Figure 2A and 2B). vSMC1 was enriched for chromatid-related processes, vSMC2 displayed a typical contractile phenotype, with enrichment for oxidative phosphorylation pathways, and vSMC3 was enriched for RNA processing pathways. vSMC4 exhibited an extracellular matrix-producing phenotype characteristic of synthetic vSMCs^22^, whereas vSMC5 was characterized by reduced expression of canonical vSMC markers and enrichment for ciliary and immune-related genes. Similarly, five subpopulations of PVFs were identified (PVF1-5, Figure 2C and 2D). PVF1 exhibited a typical fibroblast gene expression signature, whereas PVF2 was enriched for genes related to extracellular matrix production PVF3 for oxidative phosphorylation, and PVF4 for cell-cell junctions. PVF5 showed enrichment for immune-related pathways. Four subclusters were identified within the vEC population (vEC1-4; Figure 2E and 2F). However, as relatively few vECs were captured, the clusters were lesss distinct than those of the other cell types. Consequently, the gene expression patterns of vEC1, vEC2, and vEC4 displayed slight heterogeneity, which may reflect genuine biological diversity or technical noise. In contrast, vEC3 exhibited a distinct immune-related phenotype, characterized by expression of inflammation markers, such as NFKB2 and ICAM1, and enrichment for immune-related pathways.

**Figure 2:**
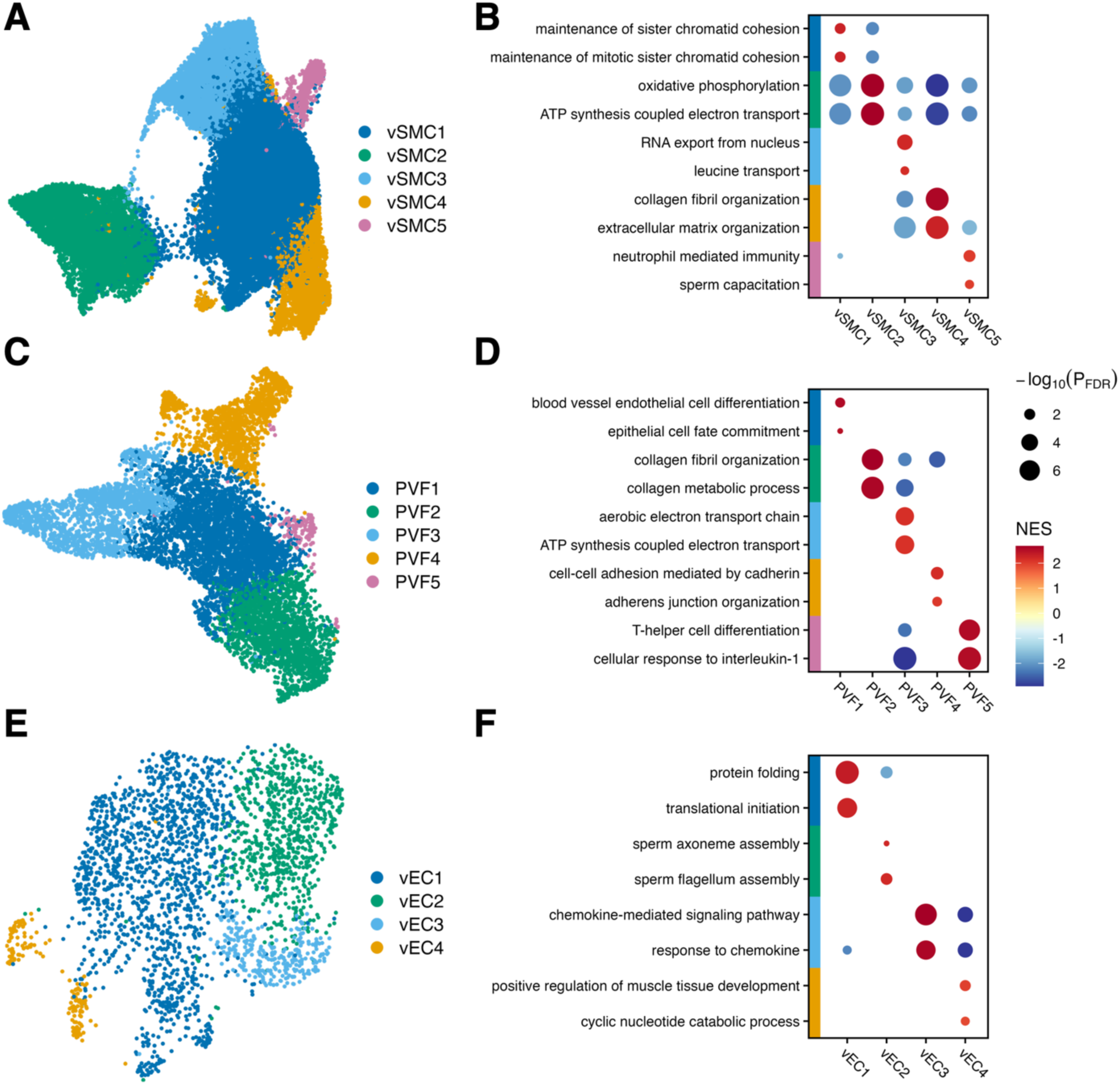
Level 2 subclustering of structural arterial cell types: Uniform Manifold Approximation and Projection (UMAP) visualizations of subclustering results for (**A**) vascular smooth muscle cells (vSMCs), (**C**) perivascular fibroblasts (PVFs), and (**E**) vascular endothelial cells (vECs). Dot plot with subclustering gene set enrichment analysis results with gene ontology biological processes for (**B**) vSMCs, (**D**) PVFs and (**F**) vECs. FDR; false discovery rate; NES, normalized enrichment score.

### Sex differences in gene expression per cell type

Differential expression analysis revealed sex differences in gene expression patterns in level 1 vSMCs, PVFs and vECs (Figure 3A, 3C and 3E; Table S6), while gene set enrichment analysis identified multiple gene sets enriched in male or female samples (Fig. 3B, 3D and 3F; Table S7). The gene sets MYC targets and mTORC1 signaling, both implicated in the regulation of metabolism and proliferation^23^, were upregulated in female samples across all structural arterial cell types. The epithelial to mesenchymal transition gene set was enriched in both female vSMCs (FDR-adjusted P = 1.07×10^-5^) and PVFs (FDR-adjusted P = 2.5×10^-9^). Additionally, inflammatory response genes were enriched in female PVFs (FDR-adjusted-P = 5.8×10^-8^), while other inflammatory gene sets, such as TNFα signalling via NF-κB, were enriched in male vECs (FDR-adjusted P = 5.0×10^-9^) and female PVFs (FDR-adjusted P = 2.5×10^-9^).

**Figure 3:**
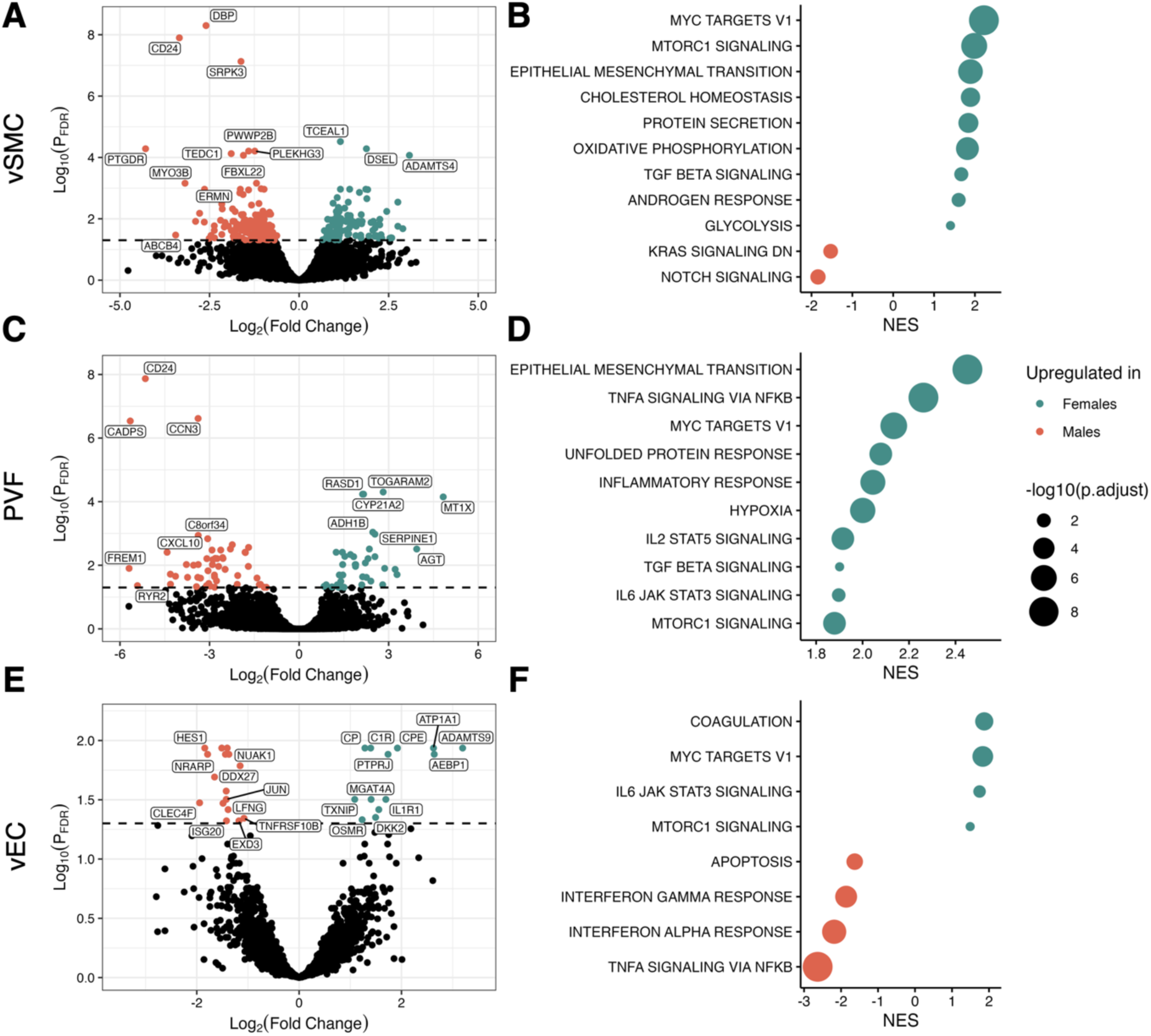
Gene expression profiles in structural arterial cell types are different between male and female samples. Volcano plots of differentially expressed genes in (**A**) vascular smooth muscle cells (vSMCs), (**C**) perivascular fibroblasts (PVFs), and (**E**) vascular endothelial cells (vECs). Dot plot of GSEA results with hallmark gene sets for (**B**) vSMCs, (**D**) PVFs, and (**F**) vECs. Only the first statistically significant gene sets are shown. FDR; false discovery rate; NES, normalized enrichment score.

### Sex differences in cell type proportions

In both level 1 and level 2 clustering analyses, we found no statistically significant differences in cell type proportions between sexes (Table S8). Also, after performing bulk RNA-seq deconvolution of IA samples using our atlas as a reference (Table S9), no statistically significant differences in cell type proportions in IAs were observed between male and female patients (Figure 4A and Table S10). This finding persisted when the analysis was restricted to structural cell types (Figure 4B). In contrast, we observed a statistically significant increase in the proportion of myeloid lineage cells in ruptured IAs compared to unruptured IAs (mean proportion unruptured = 0.088, mean proportion ruptured = 0.248, FDR-adjusted P-value = 0.0095) (Figure 4C and 4D; Table S11).

**Figure 4:**
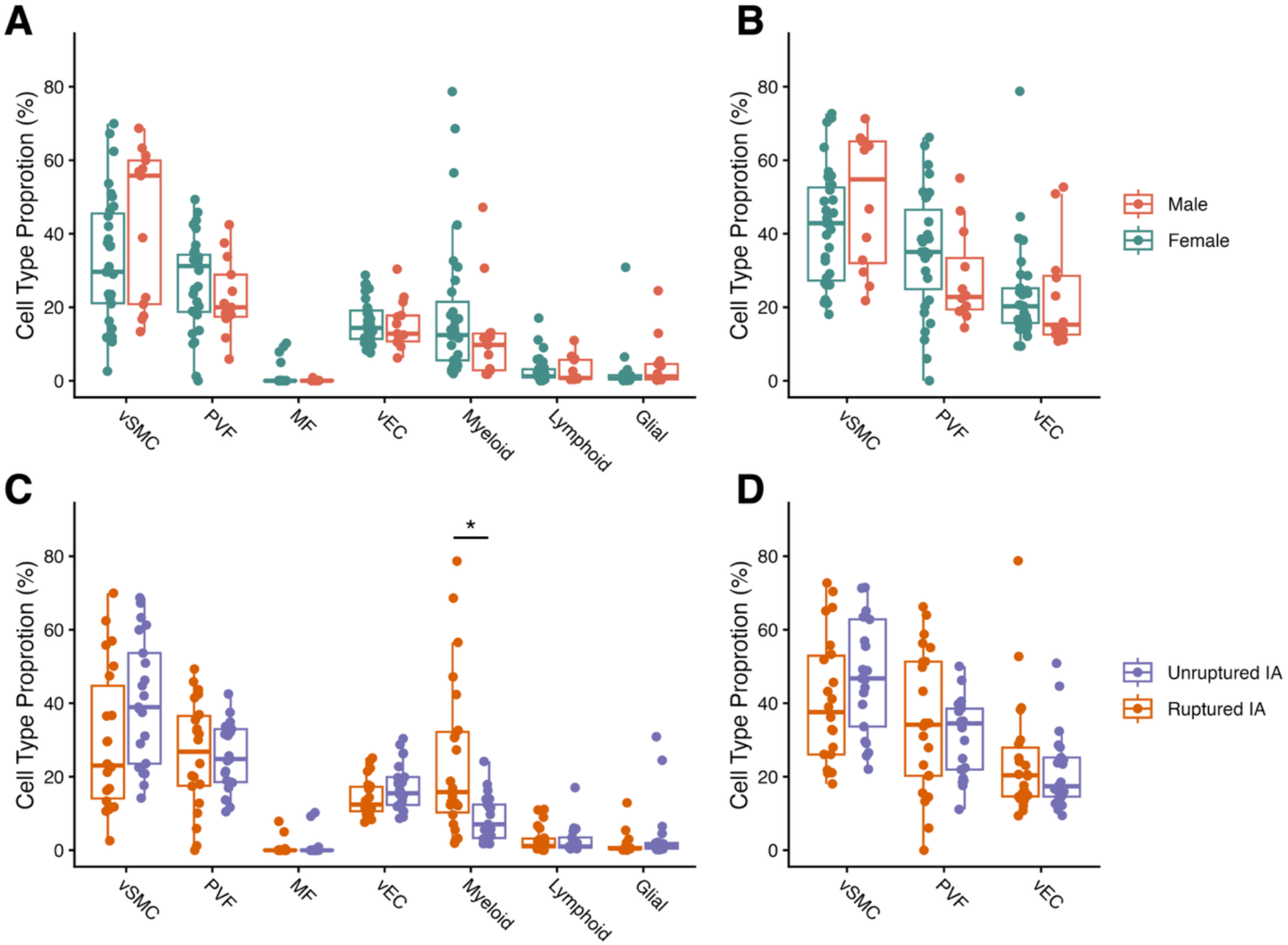
Cell type proportions from deconvolution of intracranial aneurysm bulk RNA sequencing samples. Cell type proportions from bulk RNA sequencing data^9^ were acquired though deconvolution analysis. Boxplots of the proportions of all cell types, split by (**A**) sex and (**C**) rupture status. Boxplots of the proportions of structural arterial cell types, split by (**B**) sex and (**D**) rupture status (* = false discovery rate adjusted P-value < 0.05). MF, meningeal fibroblast; PVF, perivascular fibroblast; vEC, vascular endothelial cell; vSMC, vascular smooth muscle cell.

## Discussion

In this study, we present the first scRNA-seq atlas of intracranial arteries of the CoW, encompassing 55,371 cells. Using level 1 clustering, we identified seven major cell types, while the more detailed level 2 clustering revealed 14 subpopulations within the primary structural arterial cell types, vSMCs, PVFs, and vECs. Our analyses highlighted pronounced sex-specific differences in gene expression across multiple key biological pathways, despite no detectable sex differences in overall cell type proportions in either our scRNA-seq dataset or deconvoluted bulk RNA-seq data from IA samples^9^. Additionally, we observed a higher abundance of myeloid lineage cells in ruptured IAs compared to unruptured IAs.

No previous scRNA-seq datasets of intracranial arterial tissue are available for direct comparison. Therefore, we compared our results with scRNA-seq datasets of related vascular tissues^6,24–26^. Overall, the major cell types we identified are largely consistent with these studies, although some distinct cell populations have been reported, likely reflecting differences in sampling strategies or biological context. For example, the mouse scRNA-seq dataset of IA and control intracranial artery samples reported a greater number of immune cells with more granular subpopulations^24^. This may reflect the comparison of healthy CoW arteries in our study, with potentially inflamed IAs, containing a broader diversity of immune cell states. Interspecies differences in cell composition may also contribute to this discrepancy. Pericytes were detected in human scRNA-seq datasets derived from arteries of the hippocampus and cortex^25^ and non-cranial arteries^26^, likely due to the inclusion of smaller blood vessels, such as capillaries, where pericytes are typically located. In contrast, our dataset focused on large arteries of the CoW, where pericytes are generally absent.^27^ This is consistent with observations in a scRNA-seq study of superficial temporal arteries and IAs.^6^ Finally, the presence of MFs in our dataset, also reported in a distal artery dataset^25^, may reflect by the proximity of our sampling site near the subarachnoid membrane.

We identified multiple molecular pathways that were differentially regulated between male and female arterial CoW samples. Notably, genes in the epithelial-to-mesenchymal transition (EMT) pathway were upregulated in female vSMCs and PVFs compared to males. EMT encompasses several transitional mechanisms, including endothelial-to-mesenchymal transition (EndoMT), which plays a key role in the pathophysiology of cardiovascular disease^28^, including extracranial aneurysm, such as aortic aneurysms.^29^ Interestingly, a study reported higher EMT gene expression in male coronary arteries and several other tissues^30^, whereas our data show upregulation in females, suggesting that EMT/EndoMT expression may be differentially regulated across arterial beds. In the context of IAs, emerging evidence implicates EMT/EndoMT in disease development. For example, differentially expression of EMT genes has been reported between healthy artery samples, unruptured IAs and ruptured IAs.^31^ Future studies should determine whether EndoMT, or EMT in general, contributes to the observed sex differences in IA pathogenesis.

In addition to EMT-related pathways, several other signaling pathways exhibited sex-specific gene expression. Genes associated with MYC targets and mTORC1 signaling pathways were consistently upregulated across all female structural arterial cell types, suggesting a pervasive sex-specific effect. In contrast, genes in the TNFα signaling via NF-κB pathway were upregulated in female PVFs but downregulated in female vECs. Together, these findings indicate substantial sex-specific regulation of arterial cells. Whether these pathways also contribute to the observed sex differences in IA pathogenesis remains to be determined.

We found that ruptured IAs contained a higher proportion of myeloid lineage cells, which is consistent with previous reports. For example, a histological study of six ruptured and six unruptured IAs demonstrated greater macrophages and leukocytes infiltration in ruptured compared to unruptured IAs.^32^ These findings may suggest that IAs with increased myeloid cell infiltration may have an increased risk of rupture. However, in both bulk RNA and histological analyses, it is not possible to distinguish myeloid cells present prior to rupture from those that infiltrate afterward. Therefore, we cannot conclude whether an elevated proportion of myeloid cells is a cause or a consequence of rupture, and future studies, are needed to address this question.

The main strength of this dataset lies in its high-quality gene expression profiles of a large number of cells with clear separation among the included cell types. However, our study also has certain limitations that should be acknowledged. First, we used complete CoW arteries to construct a representative scRNA-seq atlas, and, as a result, more of the abundant cell type vSMCs were sequenced than, for example, vECs. The underrepresentation of specific cell types may limit the utility of the atlas for studies focusing on rarer cell types, as it can make identifying disease-relevant subpopulations more difficult. This limitation could be mitigated in future scRNA-seq studies by enriching for these less abundant cell types. Second, we observed substantial variation in IA cell type proportions in our RNA deconvolution analysis. Given the pronounced structural heterogeneity of IA tissues, both within a single IA and across IAs^33^, the sample area selection may introduce variability in the observed cell type proportions. This variation could, in turn, limit the statistical power to detect differences between groups.

In conclusion, we present a publicly available single-cell atlas of the human intracranial arteries of the CoW. Our analyses revealed widespread cell type-specific differences in gene regulation, which may contribute to sex differences in IA prevalence and rupture risk. This atlas provides a valuable resource for studying the cell-type-specific biology of intracranial arteries of the CoW. Future integration of our dataset with scRNA-seq data from IAs or other omics-based datasets could yield important insights into IA pathogenesis. Beyond IA research, this atlas may also facilitate the study of intracranial arterial diseases, such as arteriovenous malformations or moyamoya disease.

## Abbreviations

CoW: circle of Willis
EndoMT: endothelial-to-mesenchymal transition
EMT: epithelial-to-mesenchymal transition
IA: intracranial aneurysm
MF: meningeal fibroblast
PVF: perivascular fibroblast
scRNA-seq: single-cell RNA sequencing
UMI: unique molecular identifier
vEC: vascular endothelial cell
vSMC: vascular smooth muscle cell

## Acknowledgements

We would like to thank The Netherlands Brain Bank for providing the samples for this research and Onur Basak for providing valuable insights in single cell data analysis.

---

**Figure S1.**
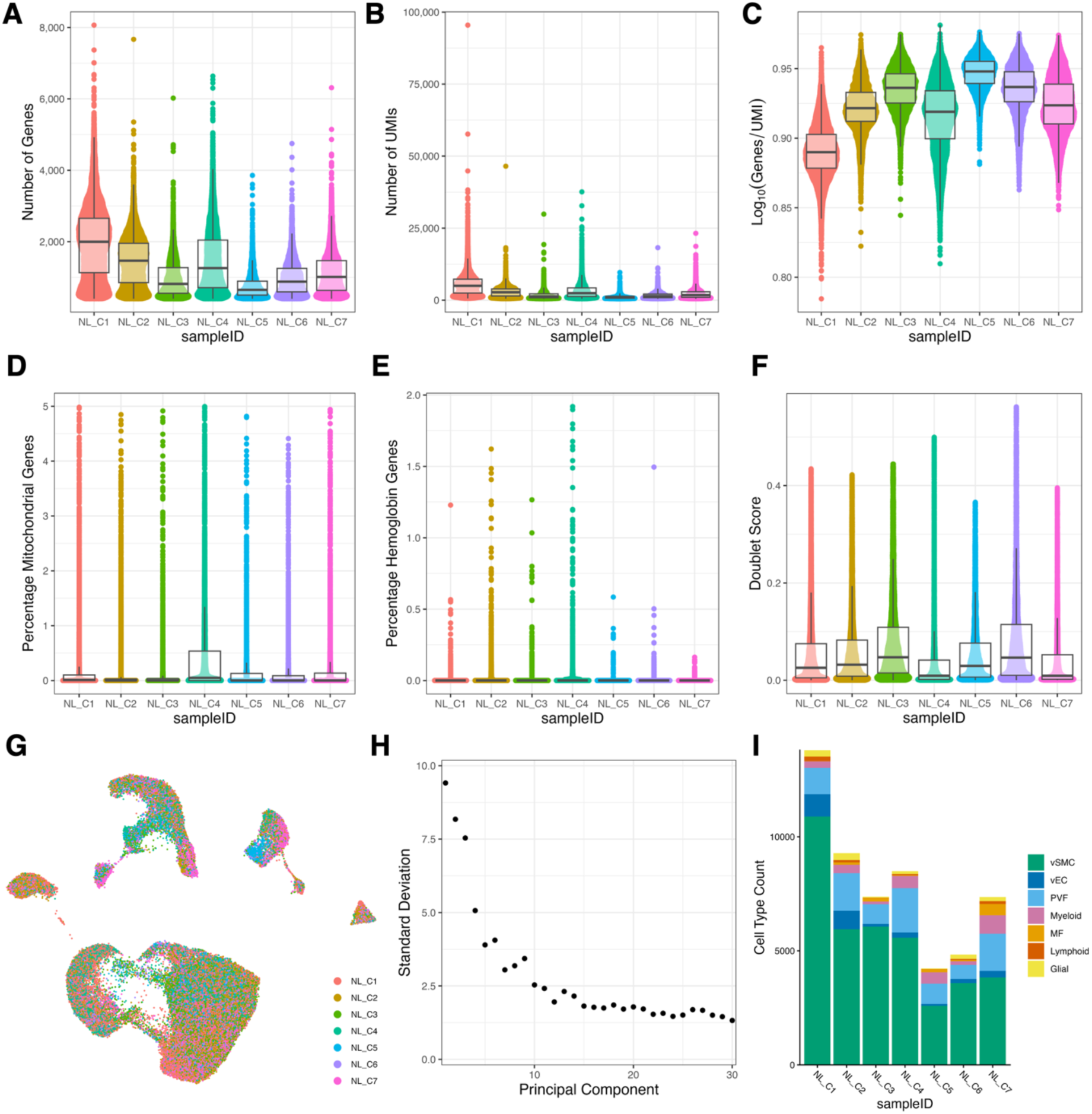
Quality control metrics of circle of Willis samples. Violin and box plots per-sample with number of genes (**A**), number of unique molecular identifiers (UMI) (**B**), log10 of genes divided by UMI (**C**), percentage mitochondrial genes (**D**), percentage of hemoglobin genes (**E**), and doublet score from the scDblFinder package (**F**). (**G**) Uniform Manifold Approximation and Projection (UMAP) colored by samples. (**H**) Elbow plot of the variance explained by the integrated principal components. (**I**) Bar graph showing cell type counts for the individual samples. MF, meningeal fibroblast; PVF, perivascular fibroblast; vEC, vascular endothelial cell; vSMC, vascular smooth muscle cell.

## References

1. Iqbal S. A Comprehensive Study of the Anatomical Variations of the Circle of Willis in Adult Human Brains. J Clin Diagn Res. 2013;7:2423–2427.

2. Vlak MH, Algra A, Brandenburg R, Rinkel GJ. Prevalence of unruptured intracranial aneurysms, with emphasis on sex, age, comorbidity, country, and time period: a systematic review and meta-analysis. The Lancet Neurology. 2011;10:626–636.

3. Nieuwkamp DJ, Setz LE, Algra A, Linn FH, Rooij NK de, Rinkel GJ. Changes in case fatality of aneurysmal subarachnoid haemorrhage over time, according to age, sex, and region: a meta-analysis. The Lancet Neurology. 2009;8:635–642.

4. Kataoka H. Molecular Mechanisms of the Formation and Progression of Intracranial Aneurysms. Neurol Med Chir (Tokyo). 2015;55:214–229.

5. Ji H, Li Y, Sun H, Chen R, Zhou R, Yang Y, Wang R, You C, Xiao A, Yi L. Decoding the Cell Atlas and Inflammatory Features of Human Intracranial Aneurysm Wall by Single-Cell RNA Sequencing. Journal of the American Heart Association. 2024;13:e032456.

6. Yu G, Li J, Zhang H, Zi H, Liu M, An Q, Qiu T, Li P, Song J, Liu P, et al. Single-cell analysis reveals the implication of vascular endothelial cell-intrinsic ANGPT2 in human intracranial aneurysm. Cardiovascular Research. 2024;cvae186.

7. Wen D, Wang X, Chen R, Li H, Zheng J, Fu W, Zhang T, Yang M, You C, Ma L. Single-Cell RNA Sequencing Reveals the Pathogenic Relevance of Intracranial Atherosclerosis in Blood Blister-Like Aneurysms. Front Immunol. 2022;13:927125.

8. Lee RM. Morphology of cerebral arteries. Pharmacol Ther. 1995;66:149–173.

9. Kleinloog R, Verweij BH, van der Vlies P, Deelen P, Swertz MA, de Muynck L, Van Damme P, Giuliani F, Regli L, van der Zwan A, et al. RNA Sequencing Analysis of Intracranial Aneurysm Walls Reveals Involvement of Lysosomes and Immunoglobulins in Rupture. Stroke. 2016;47:1286–1293.

10. Fleming SJ, Chaffin MD, Arduini A, Akkad A-D, Banks E, Marioni JC, Philippakis AA, Ellinor PT, Babadi M. Unsupervised removal of systematic background noise from droplet-based single-cell experiments using CellBender. Nat Methods. 2023;20:1323–1335.

11. Hao Y, Stuart T, Kowalski MH, Choudhary S, Hoffman P, Hartman A, Srivastava A, Molla G, Madad S, Fernandez-Granda C, et al. Dictionary learning for integrative, multimodal and scalable single-cell analysis. Nat Biotechnol. 2024;42:293–304.

12. Germain P-L, Lun A, Garcia Meixide C, Macnair W, Robinson MD. Doublet identification in single-cell sequencing data using scDblFinder. F1000Res. 2022;10:979.

13. Traag VA, Waltman L, van Eck NJ. From Louvain to Leiden: guaranteeing well-connected communities. Sci Rep. 2019;9:5233.

14. Gábor Csárdi and Tamás Nepusz. The igraph software package for complex network research. InterJournal. 2006;Complex Systems:1695.

15. Lun A. bluster: Clustering Algorithms for Bioconductor. 2023; Available from: https://bioconductor.org/packages/bluster

16. Marsh S, Salmon M, Hoffman P. samuel-marsh/scCustomize: Version 2.1.2. 2024;

17. Phipson B, Sim CB, Porrello ER, Hewitt AW, Powell J, Oshlack A. propeller: testing for differences in cell type proportions in single cell data. Bioinformatics. 2022;38:4720–4726.

18. Love MI, Huber W, Anders S. Moderated estimation of fold change and dispersion for RNA-seq data with DESeq2. Genome Biology. 2014;15:550.

19. Xu S, Hu E, Cai Y, Xie Z, Luo X, Zhan L, Tang W, Wang Q, Liu B, Wang R, et al. Using clusterProfiler to characterize multiomics data. Nat Protoc. 2024;19:3292– 3320.

20. Liberzon A, Birger C, Thorvaldsdóttir H, Ghandi M, Mesirov JP, Tamayo P. The Molecular Signatures Database (MSigDB) hallmark gene set collection. Cell Syst. 2015;1:417–425.

21. Chu T, Wang Z, Pe’er D, Danko CG. Cell type and gene expression deconvolution with BayesPrism enables Bayesian integrative analysis across bulk and single-cell RNA sequencing in oncology. Nat Cancer. 2022;3:505–517.

22. Rensen SSM, Doevendans PAFM, van Eys GJJM. Regulation and characteristics of vascular smooth muscle cell phenotypic diversity. Neth Heart J. 2007;15:100–108.

23. Pourdehnad M, Truitt ML, Siddiqi IN, Ducker GS, Shokat KM, Ruggero D. Myc and mTOR converge on a common node in protein synthesis control that confers synthetic lethality in Myc-driven cancers. Proceedings of the National Academy of Sciences. 2013;110:11988–11993.

24. Martinez AN, Tortelote GG, Pascale CL, McCormack IG, Nordham KD, Suder NJ, Couldwell MW, Dumont AS. Single-Cell Transcriptome Analysis of the Circle of Willis in a Mouse Cerebral Aneurysm Model. Stroke. 2022;53:2647–2657.

25. Yang AC, Vest RT, Kern F, Lee DP, Agam M, Maat CA, Losada PM, Chen MB, Schaum N, Khoury N, et al. A human brain vascular atlas reveals diverse mediators of Alzheimer’s risk. Nature. 2022;603:885–892.

26. Zhao Q, Pedroza A, Sharma D, Gu W, Dalal A, Weldy C, Jackson W, Li DY, Ryan Y, Nguyen T, et al. A cell and transcriptome atlas of human arterial vasculature. Cell Genomics. 2025;101034.

27. Armulik A, Genové G, Betsholtz C. Pericytes: Developmental, Physiological, and Pathological Perspectives, Problems, and Promises. Developmental Cell. 2011;21:193–215.

28. Kovacic JC, Dimmeler S, Harvey RP, Finkel T, Aikawa E, Krenning G, Baker A. Endothelial to Mesenchymal Transition in Cardiovascular Disease. J Am Coll Cardiol. 2019;73:190–209.

29. Millar JK, Salmon M, Nasser E, Malik S, Kolli P, Lu G, Pinteaux E, Hawkins RB, Ailawadi G. Endothelial to mesenchymal transition in the interleukin-1 pathway during aortic aneurysm formation. J Thorac Cardiovasc Surg. 2024;167:e146–e158.

30. Hartman RJG, Mokry M, Pasterkamp G, den Ruijter HM. Sex-dependent gene co-expression in the human body. Sci Rep. 2021;11:18758.

31. Jiang Y, Leng J, Lin Q, Zhou F. Epithelial–mesenchymal transition related genes in unruptured aneurysms identified through weighted gene coexpression network analysis. Sci Rep. 2022;12:225.

32. Korkmaz E, Kleinloog R, Verweij BH, Allijn IE, Hekking LHP, Regli L, Rinkel GJE, Ruigrok YM, Andries Post J. Comparative Ultrastructural and Stereological Analyses of Unruptured and Ruptured Saccular Intracranial Aneurysms. J Neuropathol Exp Neurol. 2017;76:908–916.

33. Wälchli T, Ndengera M, Constanthin PE, Bisschop J, Morel S, Gautschi O, Berhouma M, Kalyvas A, Monnier PP, Winkler EA, et al. Sex-Dependent Manifestations of Intracranial Aneurysms. Stroke: Vascular and Interventional Neurology. 2024;4:e001091.

34. Maas DP. Created in BioRender [Internet]. 2025. Available from: https://BioRender.com/xy1slpz

